# Initial Experience with Intercostal Insertion of an Extravascular ICD Lead Compatible with Existing Pulse Generators

**DOI:** 10.1101/2023.02.21.23286264

**Authors:** Martin C. Burke, Reinoud E. Knops, Vivek Reddy, Johan Aasbo, Michael Husby, Alan Marcovecchio, Mark O’Connor, Rick Sanghera, Don Scheck, Shari Pepplinkhuizen, Adrian Ebner

**Affiliations:** CorVita Science Foundation, Chicago, Illinois; Department of Clinical and Experimental Cardiology, Amsterdam UMC, Academic Medical Center, Amsterdam, the Netherlands; Mount Sinai Medical School, New York, NY; Lexington Cardiology and Baptist Health, Lexington, KY; AtaCor Medical, Inc., San Clemente, California; Cardiovascular Department, Sanatorio Italiano, Asunción, Paraguay

**Author notes:** Address for correspondence: Martin C. Burke, DO, CorVita Science Foundation, 1006 S Michigan Ave, Suite 500, Chicago, IL 60605, United States. Presented in part as a Late Breaking Clinical Trial at the 2022 Asia-Pacific Heart Rhythm Society Meeting, Singapore, Singapore. **Tweet:** Early experience inserting a novel, protype ICD lead via the anterior intercostal left parasternal approach into the anterior mediastinum outside the myocardium is feasible anatomically as well as functionally using commercial ICD pulse generators acutely and after 90 days. Extravascular#ICD#sudden cardiac death.

**Keywords:** ICD, extravascular, defibrillation, anterior mediastinum, intercostal

## Abstract

**Objectives:** This study assessed safety and feasibility of a novel extravascular (EV) implantable cardioverter-defibrillator (ICD) lead when inserted anteriorly through a rib space and connected to various commercially available ICD pulse generators (PGs) placed in either a left mid-axillary or left pectoral pocket.

**Background:** Currently available or investigational, EV-ICDs include a subcutaneous or subxiphoid lead connected to customized EV-ICD PGs. This novel EV-ICD (AtaCor Medical, Inc, San Clemente, CA) employs a unique intercostal implant technique and is designed to function with commercial DF-4 ICD PGs.

**Methods:** In this non-randomized, single-center, acute study, 36 de novo or replacement transvenous (TV) ICD patients enrolled to receive a concomitant EV-ICD lead inserted through an intercostal space along the left parasternal margin. EV-ICD leads were connected to DF-4 compatible ICD PGs positioned in either a left mid-axillary or pectoral pocket for acute sensing and defibrillation testing. Defibrillation testing started at 30 Joules (J) and stepped down in 10 J increments following conversion success and stepped up in 5 J increments following conversion failure.

**Results:** Successful acute defibrillation using ≤ 35 J was noted in 100% of left mid-axillary PG subjects (n=27, mean 16.3 ± 8.6 J) and 83% of left pectoral PG subjects (n=6, mean 21.0 ± 8.4 J). All evaluable episodes (n=93) were automatically sensed, detected, and shocked. No serious device-related intraoperative adverse events were observed.

**Conclusions:** This first-in-human study documented safe and reliable placement of a novel extravascular ICD lead with effective sensing and defibrillation of induced ventricular fibrillation using commercial DF-4 ICD pulse generators.

**Condensed Abstract:** This study assessed feasibility of intercostal implantation of a novel extravascular implantable cardioverter-defibrillator (ICD) lead designed to function with commercial DF-4 pulse generators (PGs). Lead placement was successful in 33 of 36 attempts (94%). Acute defibrillation with ≤35 J was successful in 27 of 27 left mid-axillary PG subjects (100%) and 5 of 6 left pectoral PG subjects (83%). All evaluable episodes (n=93) were automatically detected. No serious device-related intraoperative adverse events were observed. This study demonstrates feasibility of a novel extravascular ICD lead with effective sensing and defibrillating of induced ventricular fibrillation using commercial DF-4 PGs.

## Introduction

The implantable cardioverter-defibrillator (ICD) is widely used for primary and secondary prevention of sudden cardiac death (SCD) among patients who are at high risk for ventricular tachyarrhythmia.^1, 2^ The mortality benefit conferred with transvenous ICDs (TV-ICDs) is not without risk of acute lead complications, including hemothorax, pneumothorax, cardiac perforation, and tamponade^3-7^ and delayed complications, which consist primarily of lead failures, venous thrombosis, and bloodstream infection.^8-13^ In certain sub-populations, such as end-stage renal disease patients requiring dialysis, the risks of bloodstream infection and/or central venous stenosis outweigh potential benefit of transvenous ICD implantation despite high risk of SCD.^14^ Additionally, transvenous ICD lead placement may be precluded in patients with occluded or limited venous access, prior thoracic radiotherapy, or anomalous cardiac anatomy.^15,16^ These shortcomings have spurred development of alternative approaches that avoid vascular access and indwelling intravascular leads.^15,17,18^

Extravascular ICD (EV-ICD) systems provide an alternative to the transvenous ICDs in patients without the need for bradycardia pacing. The commercially available subcutaneous ICD (S-ICD; Boston Scientific, Marlborough, MA) has been shown to have low complication rates^19,20^ and similar efficacy to the TV-ICD, but has no pacing capability and requires a unique ICD PG capable of delivering higher energy shocks (80 J) and is, therefore, larger than modern TV-ICDs. Placement of extravascular defibrillation electrodes in closer proximity to the heart requires less shock energy for defibrillation, and therefore smaller pulse generators, compared with the S-ICD.^21,22,23^ To that end, substernal EV-ICD leads (Medtronic Inc, Minneapolis, MN) have been developed. Crozier et al.^24^reported successful substernal implantation in 299 of 316 subjects (95%) with no procedural complications observed. Shock termination of induced ventricular arrhythmias was successful in 298 of 302 subjects (98.7%) when connected to a custom EV-ICD pulse generator (Medtronic, Inc, Minneapolis, MN) placed into a left posterior-lateral pocket.

This report describes an acute, first-in-human use of a different EV-ICD lead system that is implanted via an anterior chest approach and connected to standard energy, commercially available DF-4 ICD models.

## Methods

Two non-randomized, first-in-human studies were conducted at a single tertiary hospital in Asuncion, Paraguay: (Phase I) The Parasternal Access for Shocks and Pacing with an Acutely Placed Less-Invasive Lead for EV-ICD (PASS PULL EV ICD) acute study [NCT05099289] and (Phase II) the Sensing and Defibrillation with a Commercially Available ICD Coupled With a Parasternal Extravascular Lead (SECURE EV) acute + 90-day Study [NCT05352776]. The studies were approved by the ethics committee and complied with Paraguayan national regulations and the Declaration of Helsinki. All subjects gave informed consent prior to study enrollment.

Acute implant procedure and defibrillation testing data from Phase I and Phase II. In both phases, the study populations and implant procedure methods are the same. A stronger understanding of the proof of concept (Phase I) and pilot (Phase II) using commercial ICDs for defibrillation threshold testing is presented here using the matched methods. This study will also present acute adverse events resulting from the implant procedure as well as initial chronic induced VF defibrillation performance. Phase II chronic adverse events and sensing matched data require more monitoring and adjudication and will not be presented here.

### Phase I and II Study Population

Eligible subjects included adult patients at least 18 years old who were scheduled to undergo de novo or replacement ICD procedures. Key exclusions from the study included: subjects with known structural abnormalities of the thorax; subjects with a prior sternotomy or prior surgery disrupting the pericardium or tissue outside the pericardium; prior chest radiation, pericardial disease or mediastinitis; severe lung disease or known obstruction of the intended insertion location; and NYHA function classification IV.

### EV-ICD Delivery & Lead System

The EV-ICD Lead System (AtaCor Medical, Inc.; San Clemente, CA) is designed for insertion of an extravascular defibrillator lead into the anterior mediastinum through an intercostal space along the left sternal margin directly on top of the pericardium. The EV-ICD lead has 8 cm of defibrillation coil, which is divided between two electrically connected coil segments (**Figure 1, Central Illustration**). The pace/sense electrodes allow various options for pace/sense vectors, including dedicated bipolar (near field, between the two pace/sense electrodes), integrated bipolar (between either pace/sense electrode and the defibrillation coil) and far-field (from the pace/sense electrodes to the pulse generator). The DF-4 standard lead pin is designed to connect to various commercially available DF-4 compatible ICDs. The delivery tool is specific to the EV-ICD lead allowing for symmetric delivery of the coils above the heart.

**Figure 1:**
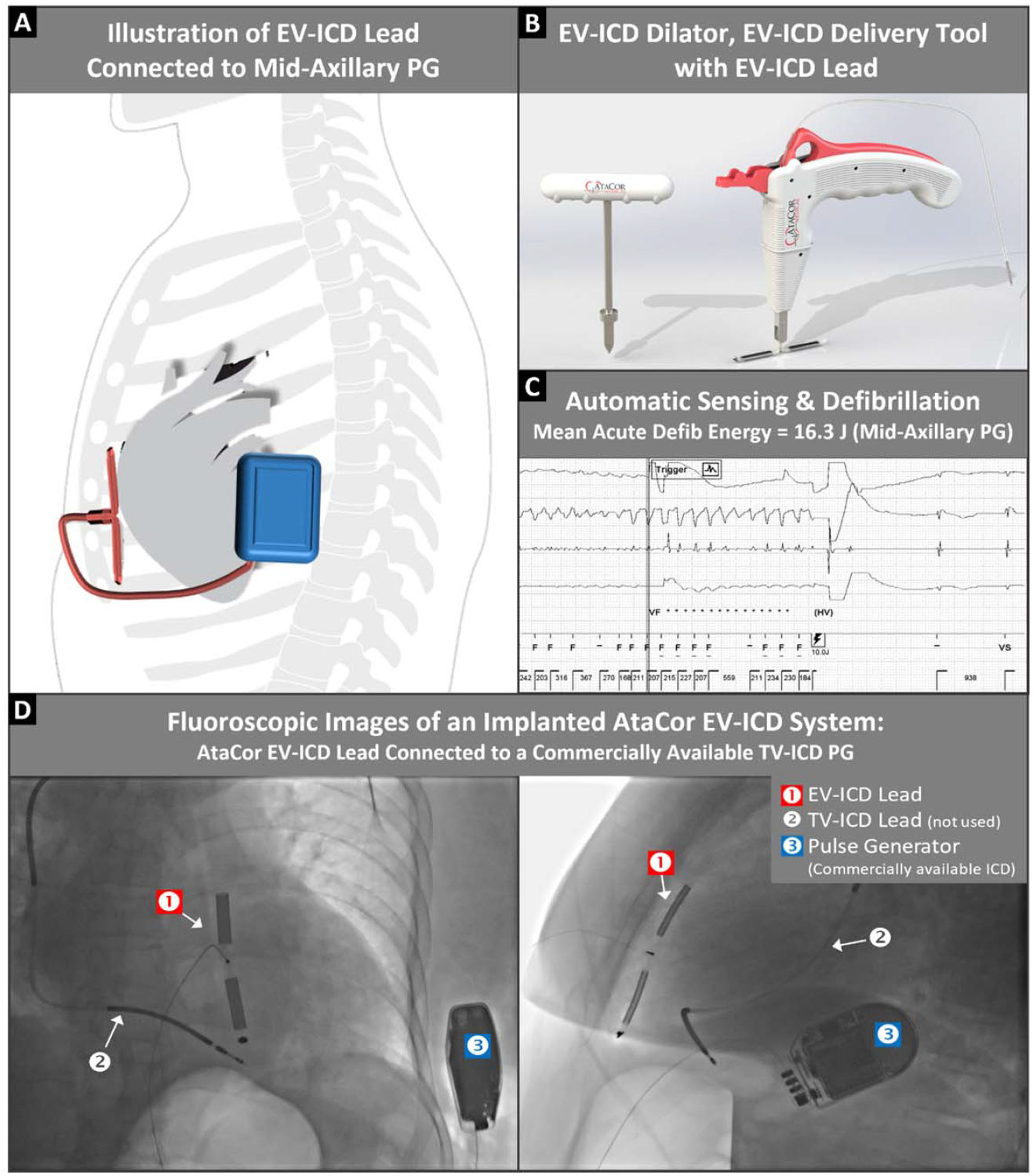
AtaCor Extravascular Implantable Cardioverter-Defibrillator (EV-ICD) Delivery and Lead System (Central Illustration). Panel A shows an illustration of the shock vector created by the implanted EV-ICD system as seen in a left lateral view. Panel B shows the EV-ICD Dilator, EV-ICD Delivery Tool and EV-ICD Lead with symmetrical, electrically connected shocking coils and pace/sense electrodes. Panel C shows automatic sensing and defibrillation of induced ventricular fibrillation with a 10 J shock. Panel D shows anterior and lateral fluoroscopic images of an EV-ICD lead connected to a left lateral DF-4 pulse generator (Abbott) and the permanent transvenous lead. PG=pulse generator; TV= transvenous; J=Joules; EV-ICD= extravascular implantable cardioverter-defibrillator.

### Implant Procedure

All implant procedures are performed under general anesthesia in a hybrid cardiac suite by experienced electrophysiologists with prior didactic and hands-on training and overseen by a cardiovascular surgeon. Surgical preparation of subjects includes placement of external defibrillator pads, standard ECG electrodes and an arterial line. **Figure 2** illustrates key aspects of the implant procedure (Phase I and II) including the anatomic target, which is typically the 4^th^ to 6^th^ intercostal space adjacent to the left sternal margin, an area that typically resides over the “cardiac notch” of the left lung. While in normal anatomy and situs the right lung extends very close to the sternal margin, the left lung does not typically extend to the left sternal margin of the inferior portion of the left chest and the cardiac notch allows for the right and left heart to reside and function un-impeded by the left lung. We have previously reported our computed tomography evaluation of the cardiac notch and this space^25^. Fluoroscopy (Anterior-Posterior and Lateral) is essential to identify the cardiac notch and guide the lead implant.

**Figure 2:**
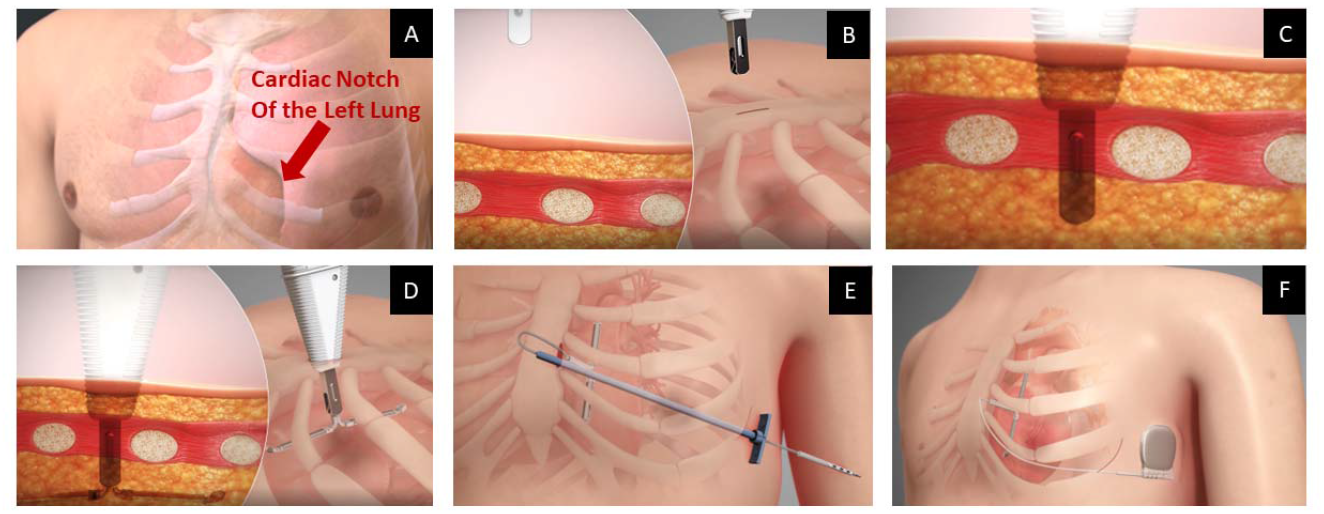
Key Aspects of the Implant Procedure. Panel A shows suitable target rib spaces at the level of the cardiac notch of the left lung. Panel B shows the approximately 2 cm incision superficial to an anterior target rib space. Panel C shows insertion of the Delivery Tool through the pathway previously created by the Dilator (not shown). Panel D shows deployment of the EV-ICD Lead. Panel E shows insertion of the proximal lead body through a peel-away sheath that was tunneled from the left mid-axillary pocket. Panel F shows the EV-ICD Lead connected to a standard DF-4 compatible ICD pulse generator in the left mid-axillary pocket.

Prior to initiation of the EV-ICD lead implant procedure, the transvenous ICD lead was placed (de novo ICD subjects) or pulse generator removed (replacement ICD subjects) to make the transvenous ICD lead available, as needed, for backup pacing and/or induction of VF for acute testing. A post transvenous lead implant echocardiogram was performed to assess for pericardial effusion.

The EV-ICD lead implant procedure required a 2-3 cm skin incision above a suitable left parasternal intercostal space that was selected based on the fluoroscopy marking of the cardiac silhouette and the left lung field. This fluoroscopic marking of key anatomic variants allowed for landing the symmetrical coils within the substernal window created by the cardiac notch of the left lung, thus avoiding entry into the pleural cavity (Figure 2). With access to the target intercostal space, the endothoracic fascia was punctured with a needle followed by an 0.035 J-tip wire to access to the mediastinum below the ribs.

The fascia anterior to the external intercostal muscle was then dilated. With the wire retained, the ICS was dilated further with a custom EV-ICD Dilator tool (AtaCor Medical, Inc.; San Clemente, CA). The EV-ICD Dilator was designed to create a more suitable passageway into the mediastinum allowing for the EV-ICD Delivery Tool to pass the endothoracic fascia and into the anterior mediastinum. The EV-ICD Delivery Tool was pre-loaded with the EV-ICD lead. Both the EV-ICD Dilator and EV-ICD Delivery Tool employed wide stoppers to limit the insertion depth and prevent over penetration damage to the pericardium and/ or myocardium. Once the EV-ICD Delivery Tool tip was inserted within the mediastinum below the endothoracic fascia, the deployment lock was released, allowing manual depression of the deployment actuator and deployment of the distal lead segment into the tissue of the mediastinum. The EV ICD lead was delivered deliberately during 90-degree lateral fluoroscopy monitoring the lead tips as it banks off the pericardium of the beating heart monitoring carefully for cardiac ectopy. Defibrillation coil segments were designed to deploy in opposing directions while banking off the pericardium. Following deployment, the EV-ICD Delivery Tool is removed over the lead body and proximal DF-4 connector, leaving the terminal pin contacts available for attachment with a traditional pacing system analyzer (PSA).

EV-ICD lead deployment and stability again were evaluated using fluoroscopy (AP and 90 degree lateral). Time to lead deployment was recorded as the time from the sternal incision to removal of the delivery tool. Echocardiography was used before and after EV-ICD lead deployment to assess for pericardial effusions. Electrical parameters including R wave amplitude, pacing impedance, and pacing capture / muscle stimulation at 10 Volts (1.5 millisecond pulse width) were measured with a standard PSA in dedicated bipolar and integrated bipolar vectors. When pacing capture was achieved, a pacing capture threshold was obtained; however, since pacing capture was not a primary objective of the study, leads were not repositioned based on outcome of the pacing capture / muscle stimulation test. Suturing material was used to attach the proximal lead segment to the parasternal fascial tissue to prevent excessive movement of the lead during acute defibrillation testing. Procedure time was recorded as the time from the incision to the end of suturing. In Phase I, the EV-ICD lead was left outside of the body for connection to an ICD pulse generator placed in either a left mid-axillary pocket (subjects 1 to 12) or pectoral pocket (subjects 13 to 18). Patients 19-36 (Phase II) had the EV-ICD lead tunneled from the sternum to the left lateral pocket for up to 90 days of implant. In both ICD pocket locations, submuscular or intermuscular PG placement was preferred, but subcutaneous placement was allowed. The choice of PG location was made without consideration of individual patient characteristics (e.g., body habitus and/or cardiac anatomy). Because the first 12 consecutive subjects established feasibility of defibrillation from the left mid-axillary PG position, the protocol was revised to allow testing from the traditional transvenous PG location (pectoral) to determine whether that PG location may also be feasible. The choice of pulse generator pocket placement was a non-random consecutive choice in Phase I only.

Pulse generators from four manufacturers (Abbott, Biotronik, Boston Scientific and Medtronic) were used. Once connected to the EV-ICD lead, the ICD programmer was used to re-measure electrical parameters and set up for acute defibrillation testing (e.g., single zone, 170-171 BPM rate cut off).

### Defibrillation Testing

The defibrillation testing protocol was designed to ascertain the lowest successful defibrillation energy within 3 shocks (**Figure 3**). Episodes of ventricular arrhythmia were induced with either the ICD connected to the EV-ICD lead or an external energy source connected to the transvenous ICD lead. Inductions were spaced by 5 minutes. Automatic sensing and detection algorithms were relied upon for the first ICD shock unless manual commanded shocks were preferred by the investigator. The defibrillation energy requirements are reported in delivered, as opposed to stored energy, as described by each manufacturer. External rescue shocks were initiated immediately after failed ICD shocks.

**Figure 3:**
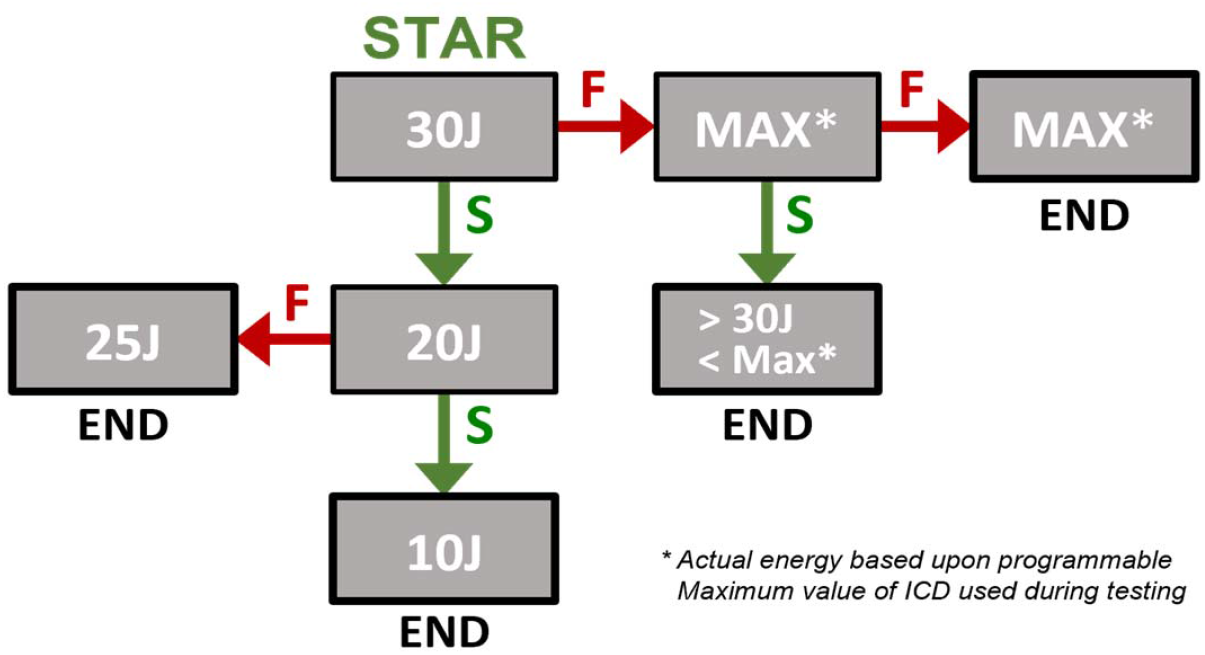
Acute Defibrillation Testing Flowchart. Sequence of shock energies tested to determine the lowest acute successful shock energy. Actual energies tested are determined by programmable options of the ICD pulse generator (for example, Boston Scientific ICDs were tested at delivered energies of 27, 18 and 10 J).

Detection time and time to therapy were measured as the time from the first sensed beat to the initiation of capacitor charging and to shock delivery, respectively. Appropriate sensing of the induced ventricular tachyarrhythmia was defined as the delivery of a shock without the need to override automatic sensing and detection algorithms with a manual shock.

### EV-ICD Lead Removal, Retention and Final Follow-Up

Following acute defibrillation testing, the study ICD pulse generator was handled in two methods. In Phase I, the study ICD was disconnected from the EV-ICD lead and connected to the transvenous lead to finish the permanent transvenous ICD implant procedure. The EV-ICD lead was then removed from the chest and a post-removal echocardiogram was performed to assess for pericardial effusions and or adverse reaction. A final follow-up was conducted 7-10 days after testing via phone, video call or inperson to document any latent adverse events associated with the study.

In Phase II (patients 19-36), the subjects retained the study extravascular ICD and a clinically indicated transvenous ICD simultaneously. The study ICD was retained in a sensing mode programmed in an identical fashion to the transvenous ICD parameters for matching which will be presented following adjudication. A post-implant echocardiogram was performed to assess for pericardial effusions and or adverse reaction. The Phase II patients will be followed comparing both devices over 90 days. Data collected beyond the procedure day (e.g., 90-day induced VF conversion, extraction experience) are not covered in this paper.

### Statistical Analysis

Descriptive statistics were used for demographics, medical history, procedure times, electrical parameters, induced episode sensing and lowest successful defibrillation testing. The study was not powered for pre-specified statistical hypothesis tests.

## Results

Thirty-six patients (18 in Phase I and 18 in Phase II) were enrolled and underwent an intercostal EV-ICD lead implant procedure between November 2021 and June 2022. The EV-ICD lead was successfully implanted in 33 of 36 subjects (92%) and 33 of 35 subjects (94%) in whom a lead insertion was attempted. Lead insertion was not attempted in one subject with intracostal ossification of the left parasternal cartilage. The two lead insertion failures include one instance where the blunt end of the EV-ICD Delivery Tool failed to penetrate the endothoracic fascia, preventing the lead from deploying in the intended location within the mediastinum. The lead was withdrawn without complication. Subsequent procedures employed an additional step to first breach the endothoracic fascia with a standard needle and introducer dilator to facilitate passage of the EV-ICD Dilator and Delivery Tool. The other case of failed lead insertion was due to mediastinal adhesions preventing full deployment of the coils. Successfully placed EV-ICD leads (n=33) were tested with a left mid-axillary PG (n=27) or left pectoral PG (n=6). The disposition of all study subjects and distribution of ICD pulse generator locations and manufacturers is shown in **Figure 4**.

**Figure 4:**
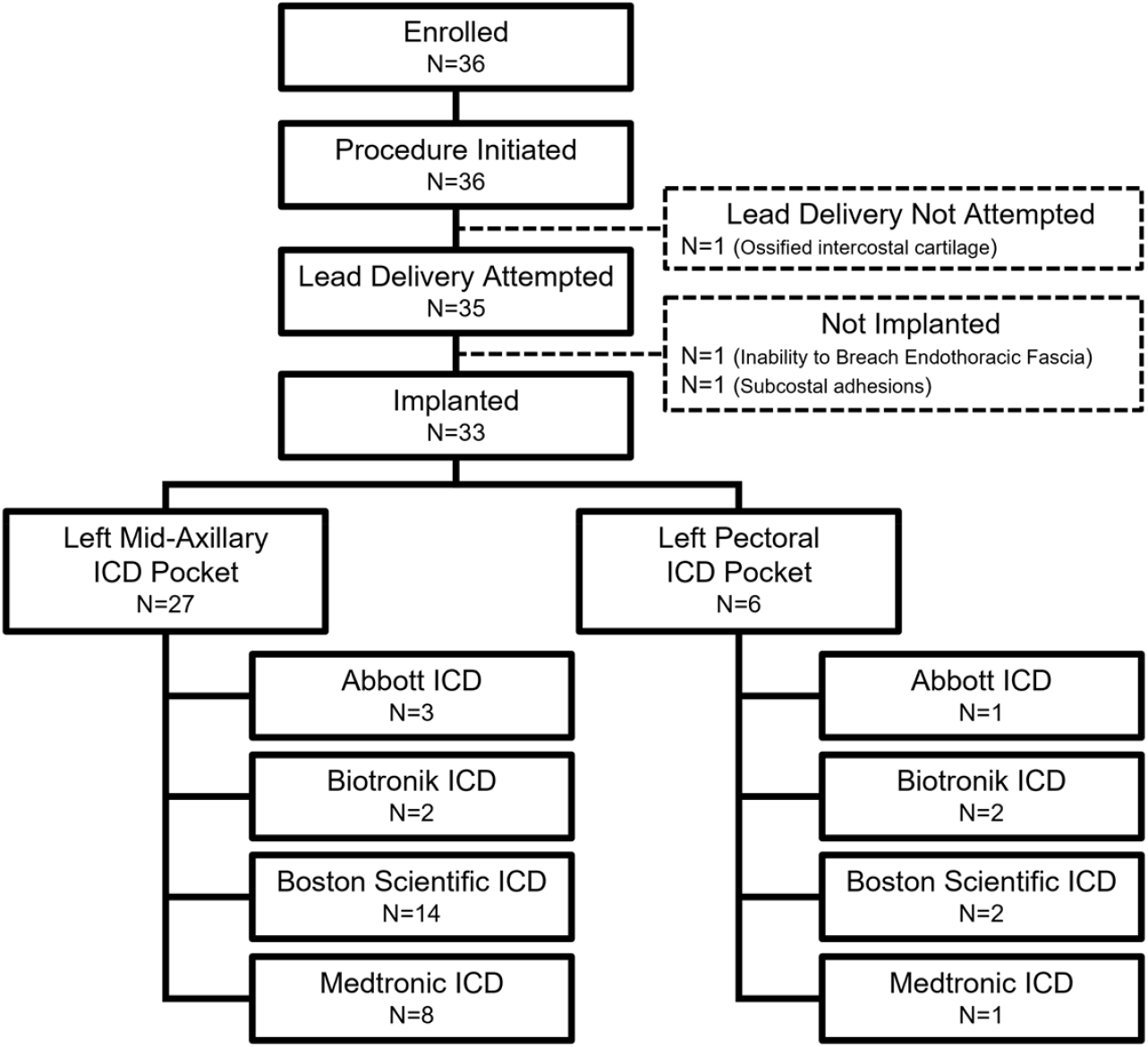
Enrollment, Implantation, and ICD Testing Summary. Disposition of all study subjects by ICD pulse generator location and manufacturer

The study cohort was predominantly male (88%) with a mean age of 58 (range 21-88) years. The mean body mass index was 28 kg/m^2^ (range 22 – 37.5). The main indication for ICD therapy was primary prevention (53%) with non-ischemic cardiomyopathy reported in 58% of subjects. Baseline characteristics are presented in **Table 1**.

**Table 1.**
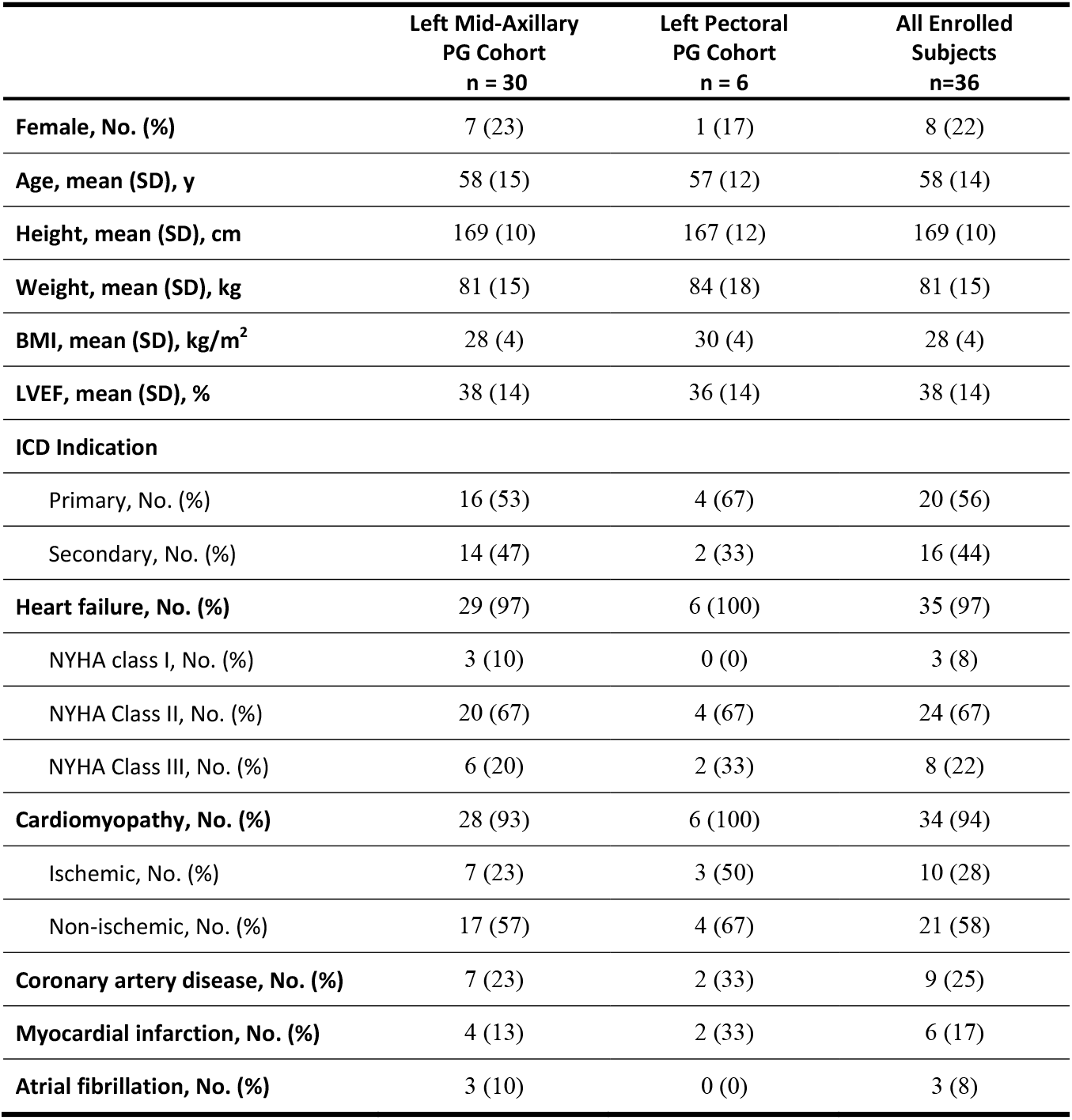
Demographics and Medical History.

|  | **Left Mid-Axillary**<br>**PG Cohort**<br>**n = 30** | **Left Pectoral**<br>**PG Cohort**<br>**n = 6** | **All Enrolled**<br>**Subjects**<br>**n=36** |
| --- | --- | --- | --- |
| **Female, No. (%)** | 7 (23) | 1 (17) | 8 (22) |
| **Age, mean (SD), y** | 58 (15) | 57 (12) | 58 (14) |
| **Height, mean (SD), cm** | 169 (10) | 167 (12) | 169 (10) |
| **Weight, mean (SD), kg** | 81 (15) | 84 (18) | 81 (15) |
| **BMI, mean (SD), kg/m<sup>2</sup>** | 28 (4) | 30 (4) | 28 (4) |
| **LVEF, mean (SD), %** | 38 (14) | 36 (14) | 38 (14) |
| **ICD Indication** |  |  |  |
| Primary, No. (%) | 16 (53) | 4 (67) | 20 (56) |
| Secondary, No. (%) | 14 (47) | 2 (33) | 16 (44) |
| **Heart failure, No. (%)** | 29 (97) | 6 (100) | 35 (97) |
| NYHA class I, No. (%) | 3 (10) | 0 (0) | 3 (8) |
| NYHA Class II, No. (%) | 20 (67) | 4 (67) | 24 (67) |
| NYHA Class III, No. (%) | 6 (20) | 2 (33) | 8 (22) |
| **Cardiomyopathy, No. (%)** | 28 (93) | 6 (100) | 34 (94) |
| Ischemic, No. (%) | 7 (23) | 3 (50) | 10 (28) |
| Non-ischemic, No. (%) | 17 (57) | 4 (67) | 21 (58) |
| **Coronary artery disease, No. (%)** | 7 (23) | 2 (33) | 9 (25) |
| **Myocardial infarction, No. (%)** | 4 (13) | 2 (33) | 6 (17) |
| **Atrial fibrillation, No. (%)** | 3 (10) | 0 (0) | 3 (8) |

There were four investigational device-related intraoperative adverse events observed within the first 7 days in both Phase I and II, including: one report of a transient pneumomediastinum, one report of atrial flutter following conversion of an induced ventricular tachyarrhythmia and one induction of ventricular tachycardia induced during the lead insertion procedure. This incidence of atrial flutter was converted to sinus rhythm with a single cardioversion shock and the incidence of ventricular tachycardia was converted with a single 200 J cardioversion shock. These three investigational device-related interoperative adverse events were resolved on the same day with no further complications. One patient in the chronic implant arm suffered an EV Lead related pneumothorax that required device removal on day 5 and a chest tube. There were no new or worsened pericardial effusions as confirmed by echocardiography post operatively, or within 7 days after the procedure.

The mean time from incision to final placement of the EV-ICD lead was 12.8 minutes (std dev 7.9; median 10.7; range 4.5, 40.3) and mean time from incision to fixation (suture) was 23.7 minutes (std dev 9.4; median 20.4; range 11.9, 56.5).

In the standard bipolar configuration (dedicated bipolar), pacing capture was observed at or below 10 volts in 6 subjects (18 %). The mean sensed R wave amplitude was 3.9 mV (SD 2.0; range 1.0, 8.1) and mean pacing impedance was 1183 ohms (SD 614; range 380, 3000). Pacing at the maximum output of the ICD and 1.5 millisecond pulse width did not produce visible or palpable muscle stimulation in any subjects.

All 33 subjects with successful EV-ICD lead placement underwent defibrillation testing. In the left mid-axillary PG cohort, 24 of 27 subjects (89%) were converted to sinus rhythm with ≤ 30 J and the mean successful defibrillation energy was 16.3 Joules (n=27; std dev 8.6; range 9, 35). All 27 of 27 subjects achieved successful VF defibrillation using ≤ 35 J (100%). In the left pectoral PG cohort, 5 of 6 subjects (83%) were converted to sinus rhythm with ≤ 35J and the mean successful defibrillation energy was 22.2 Joules (n=5; std dev 8.3; range 10, 30). In one subject, the first subject enrolled in the pectoral PG cohort, the induced episode could not be converted with 27 J or 35 J ICD shocks and required an external defibrillator shock. Subsequent procedures implemented an alternative PG placement strategy, placing it submuscular and more lateral compared to the first subject in the pectoral cohort. All subsequent subjects were successfully defibrillated with ≤ 35 J.

The mean shock impedance was 74 ohms (n=79; std dev 19; range 47-131) in shocks delivered from a left mid-axillary ICD PG and 70 ohms (n=19; std dev 5; range 62, 78) in shocks delivered from a pectoral ICD PG. The percentage of subjects successfully converted at incrementally higher energies is shown in **Figure 5**.

**Figure 5:**
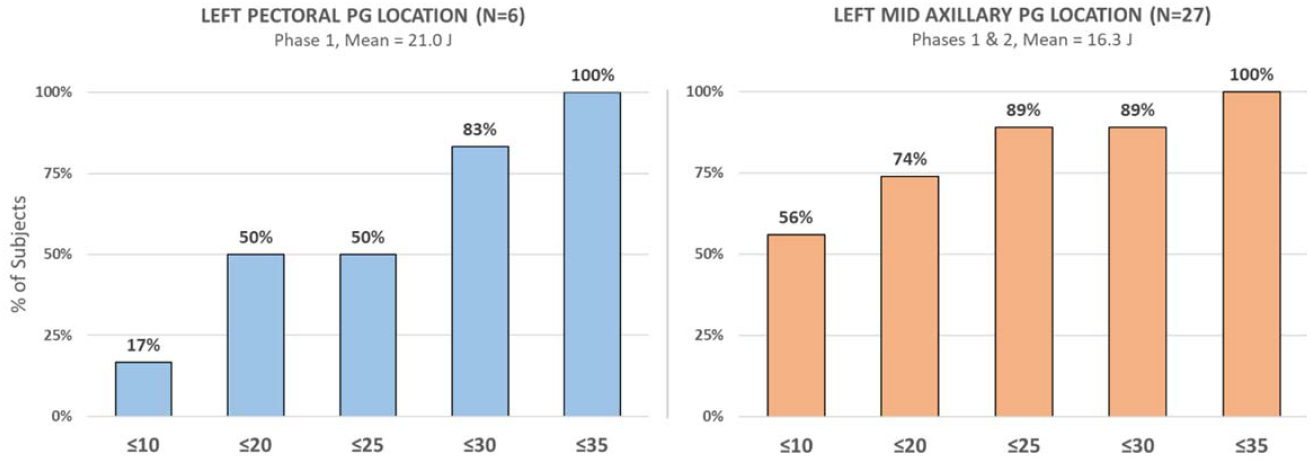
Acute Defibrillation Performance. Cumulative percentage of subjects successfully converted at incrementally higher energy levels. The single non-conversion 35 J in the left pectoral PG cohort is excluded from the mean.

VF induction testing generated a total of 98 induced episodes, in which 93 were tested using automatic detection of the ICD algorithms and five episodes controlled with manual shock delivery without automatic ICD sensing activated. Induced episodes were appropriately sensed and detected in all 98 episodes (100%) in which automatic ICD sensing was active. The four manually commanded shocks (1 at 28 J, 2 at 18 J, and 1 at 9 J) followed an automatically sensed episode in which the detection zone cutoff was inadvertently programmed to a single detection zone ≥ 250 BPM rather than the protocol-specified value of 170 BPM (recognized upon review of stored episodes following the procedure). This programming caused several fast intervals to be classified as non-VF by the device and charging was briefly interrupted. Sensing within the VF zone was reestablished without manual intervention and the episode was automatically detected and successfully terminated, albeit at the maximum 40 Joule energy, which is normal behavior of the ICD (Biotronik) after interruption of the initial charge sequence. Out of an abundance of caution, subsequent shocks were manually commanded for this patient without activation of automatic ICD sensing. As automatic ICD sensing was not evaluable in these 4 episodes, they are excluded from all sensing analyses. Undersensing was briefly noted during a single episode due to suturing that allowed the lead to be pulled back. When undersensing was noted, the implanting physician applied mild downward pressure, which resulted in normal sensing, automatic detection and shock delivery. A complete summary of detection time and time to therapy is presented in **Table 2**.

**Table 2.** Sensing of induced ventricular arrhythmias with the EV-ICD Lead.

|  | Detection Time (s) |  | Time to Therapy (s) |  |
| --- | --- | --- | --- | --- |
|  | Mean (SD) | Median [Range] | Mean (SD) | Median [Range] |
| **All Evaluable Episodes (N=93)** | 5.6 (3.4) | 4.5 [2.1, 20.1] | 10.7 (3.7) | 10.3 [4.8, 22.3] |

An example of an automatically sensed, detected, and shocked episode is shown in **Figure 6**.

**Figure 6.**
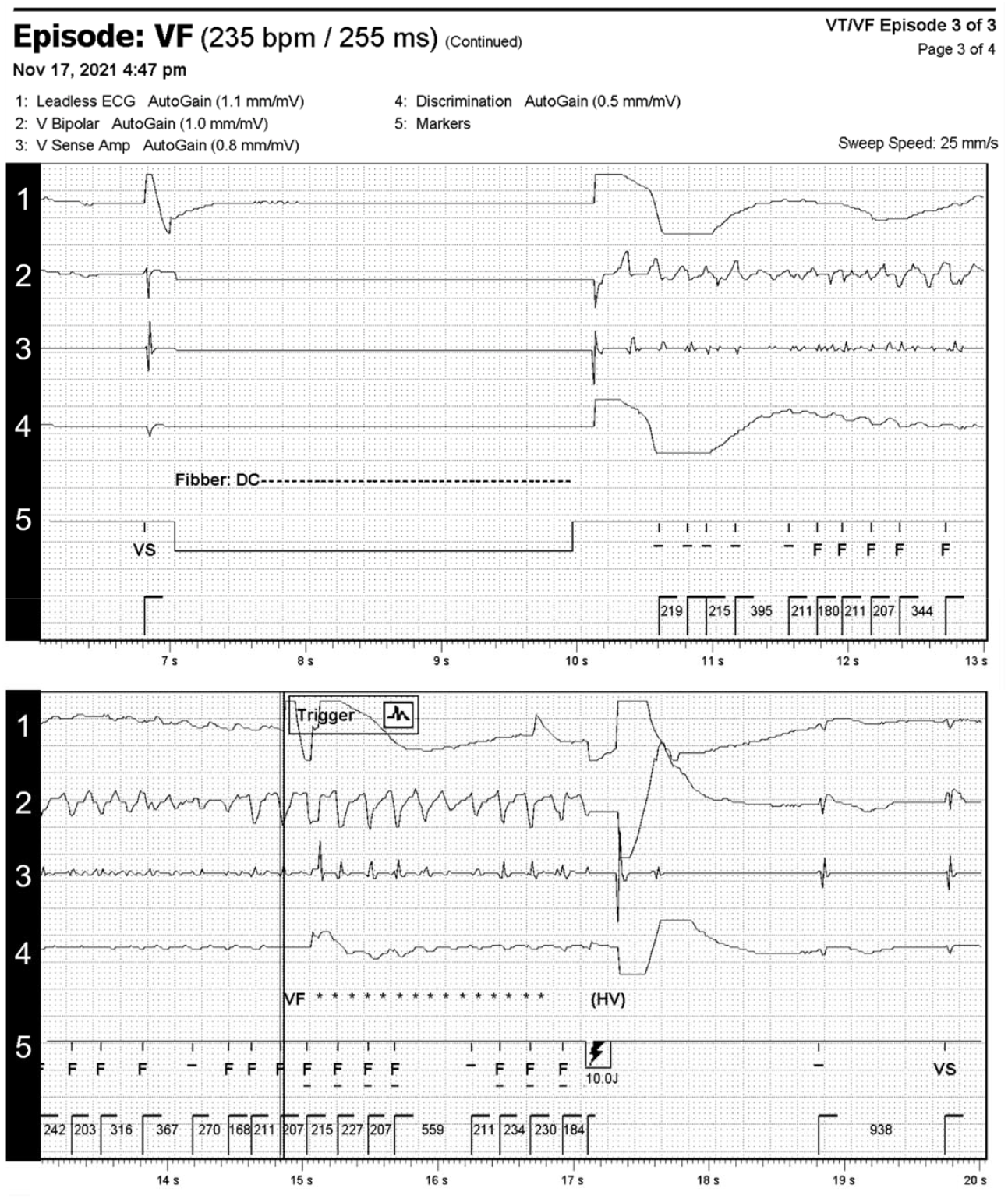
Example of sensing, detection and therapy (10 J shock) with the intercostal EV-ICD lead. Induction, sensing, detection and treatment of an induced ventricular tachyarrhythmia with the AtaCor EV-ICD Lead connected to a commercially available ICD (Abbott).

## Discussion

### Main Findings

This first-in-human, acute study demonstrates the ability to safely and reliably place an extravascular ICD lead below the sternum. This study shows that placing an EV-ICD lead in the anterior mediastinum through an intercostal space along the left parasternal margin is feasible. This EV-ICD lead, as well, can appropriately sense and defibrillate induced ventricular fibrillation using commercially available DF-4 compatible ICD pulse generators placed within a left mid-axillary or left pectoral pocket.

The high implant success rate and short lead delivery time in these first 36 subjects attest to the usability of the unique delivery tool matched to its lead system via an anterior chest approach. These metrics compare favorably with published data from other extravascular ICD systems.^22,24,26-28^ One procedure failure illuminated the challenge of penetrating the endothoracic fascia anteriorly via the intercostal access, which adheres to the posterior side of the ribs and intercostal muscles, using a blunt instrument design. Adjustments to the implant procedure were effective in overcoming this challenge and provide direction for future iterations of the procedure tools and EV-ICD lead designs. The only other failure to deliver the EV-ICD lead was noted in a patient with subcostal adhesions, which has been described previously with other systems.^22^

### Safety

In this study, the anterior insertion procedure and devices have been shown to be safe in an acute timeframe. These early results are promising if they carry forward with future operators. Similarly, Chan et al.^27^, in their acute 16 patient feasibility ASD Clinical Study, have demonstrated minimal device related complications using a subxiphoid approach including pleural damage (n=1/16; 6%) and left internal thoracic artery (LITA) damage without hemorrhage (n=1/16; 6%). However, expanding the subxiphoid approach to more centers in ASD2^28^ has reported seven adverse events in 6 patients including mortality (2/79; 3%); pericardial injury/inflammation (3/79; 4%) and hemodynamic collapse (2/79; 3%). There were no reports of left internal thoracic artery damage, although LITA without hemorrhage could have gone undetected.

The anterior approach along the parasternum can avoid the pleural space through selection of an appropriate rib space for entry. The amount of space from the sternum to the heart can also be managed from an anterior approach using delivery tool and lead designs to avoid pericardial entry or irritation. Crozier et al.^22^ in their chronic EV-ICD implant study reported some acute inspiratory discomfort but no long-term implant related pericardial inflammation or damage. Chronic data are needed to assess the safety of the deploying and maintaining a lead in the cardiac notch from an anterior intercostal approach along the left parasternum using a broader group of operators.

### Rate Sense/Pacing

The novel EV-ICD lead appropriately sensed extravascular normal sinus rhythm (R waves) and the smaller amplitude and highly variable signals seen during induced VF. Of note, VF was sensed appropriately from two different pulse generator pocket locations (left lateral & pectoral) using ICDs from multiple manufacturers, representing a variety of sensing and detection algorithms. The observed detection times and time to therapy with this new extravascular configuration have been comparable to published results from standard transvenous ICD systems with direct endocardial attachment to the myocardium.^29^

This initial EV-ICD lead design has not been optimized for cardiac pacing, while a small number of subjects achieved pacing capture intraoperatively, no subjects demonstrated pacing capture post-operatively when connected to an ICD at the maximum output. Pacing from electrodes in the extravascular substernal space has been elusive in acute and chronic studies. These studies^22,24,27,28^ have reported elevated capture thresholds, similar sensing of R wave amplitudes and chest sensation at capture outputs. Future lead iterations will be tested for their efficacy in providing bradycardia and anti-tachycardia pacing (ATP). Quast et al.^25^ have reported cardiac capture from custom designed pacing leads inserted via a similar left parasternal manner, perhaps offering a pathway for incorporation and cross engineering into future EV-ICD lead designs. Improving the rate/sense and ATP capability of the EV-CID Lead System is a major part of the future direction of research.

### Extravascular Defibrillation

Defibrillation energy requirements acutely observed in this study are similar to transvenous and substernal extravascular lead systems^22,24,27,28^, supporting effectiveness with typical safety margins using standard, commercially available DF-4 36 – 41 Joule ICD pulse generators, an important factor for pulse generator size, patient comfort, device selection among a wide range of ICD pulse generators. This study indicates that ICD pulse generators with less than 36 J maximum output (e.g., 31 J) would not provide a 10 J safety margin in some patients. The substernal extravascular configuration has tested consistently lower than the entirely subcutaneous defibrillator system despite similar position of the pulse generator in a posterior lateral pocket^22,24,27,28^. Results from this study demonstrate a 10 J safety margin may be possible in most patients using the anterior approach extravascular lead and a standard 36 J ICD placed in a left mid-axillary position or left pectoral submuscular position. Flexibility in PG pocket location allows for the use of alternate shock vectors, which may be useful for troubleshooting, and may avoid the need for an additional incision in replacement procedures. Importantly, successful defibrillation in this study was achieved with ICDs placed in submuscular device pockets; a method that is well understood with a complication profile similar to the subcutaneous method.^30^ More research is required to better understand the defibrillation performance of subcutaneously placed pulse generators in connection to this new EV-ICD Lead and position.

### Study Limitations

This study was limited as an acute assessment of sensing and defibrillation within a single center and operator in a small feasibility study population. Phase II chronic implant data including matched sensing data were not presented here due to need for external monitoring and adjudication. Longer-term data and broader clinical evaluations are required. These feasibility and pilot studies indicate that some patients implanted with an ICD pulse generator with less than 36 J maximum output (e.g., 31 J) would not have a 10 J safety margin.

## Conclusions

This first-in-human study demonstrates the safety and feasibility of sensing and defibrillating induced VF with a left parasternal extravascular defibrillator lead placed in the mediastinum through an intercostal space and connected to various DF-4 commercially available ICD pulse generators. Pacing with standard ICD outputs was not possible with this first-generation lead design. Design enhancements will be required if pacing therapies are to be made available in the future. Further data are needed to confirm chronic ICD sensing, safety performance and rhythm discrimination during ambulatory patient use.

## Data Availability

All data submitted in this manuscript is part of regulatory submissions and will be available online eventually.

## Abbreviations and Acronyms

ATP: Anti-tachycardia Pacing
EV: Extravascular
ICD: Implantable Cardioverter-Defibrillator
ICS: Intercostal Space
J: Joules
PG: Pulse Generator
S-ICD: Subcutaneous ICD
SCD: Sudden Cardiac Death
TV: Transvenous

## Acknowledgments

The authors thank study participants, Sanatorio Italiano staff (Paraguay), Sarah Hase, BS, and Kathryn Muller, PhD.

## Clinical Perspectives

a. **Clinical Competency in Patient Care and Procedural Skills:** The two prospective clinical feasibility and pilot studies presented here provide insight into a unique anatomic location and approach placing an ICD lead extravascular to the heart. Minimizing defibrillation hardware in the vasculature and heart is a contemporary topic in providing sudden cardiac death prevention to indicated patient populations. This paper has demonstrated acute procedural access, safety and efficacy of a novel ICD lead built considering the base of procedural skills of the trained electrophysiologist.
b. **Translational Outlook:** The two clinical trials, presented here, test basic research using iterative prototype defibrillation leads entering the mediastinum anteriorly. The prototypes have previously been tested in swine for defibrillation energy requirements suggesting human use with commercially available ICD pulse generators may be feasible. Our basic research data allow for the two efficient acute feasibility and pilot study designs that prove this concept true.

